# Impact of the COVID-19 pandemic on exercise habits and overweight in Japan: a nation-wide panel survey

**DOI:** 10.1101/2022.10.11.22280942

**Authors:** Sae Ochi, So Mirai, Sora Hashimoto, Yuki Hashimoto, Yoichi Sekizawa

## Abstract

**Introduction:** A catastrophic disaster may cause distant health impacts like immobility and obesity. This research aims at analysing the impact of the COVID-19 pandemic on exercise habit and overweight in the Japanese population.

**Methods:** Nation-wide online questionnaires were conducted five times from October 2020 to October 2021. The change in exercise habit, body mass index (BMI) and status of overweight (BMI>25kg/m^2^) were compared between the first questionnaire and later ones. Risk factors of losing exercise habit or developing overweight were analysed using multiple regression.

**Results:** Data was obtained from 16,642 participants. In the early phase of the pandemic, people with high income and elderly females showed higher risk of decreased exercise days. Proportion of overweight was increased from 22.2% to 26.6% in males and from 9.3% to 10.8% in females. Middle aged males, elderly females, males who experienced SARS-CoV-2 infection were at higher risks of developing overweight.

**Conclusion:** Our findings suggest that risks of immobility and overweight are homogeneous. Continuous intervention for elderly females and long-term intervention for males who were infected might be especially needed. As most disasters can cause similar social transformation, research and evaluation of immobility and obesity should be addressed in future disaster preparation/ mitigation plans.

## Introduction

During and after a catastrophic disaster population health may deteriorate in many ways. The impact on health is not limited to direct acute conditions such as injuries, but also includes indirect and chronic effects caused by lifestyle changes, mental stress, job losses, and social disruption. In particular, after chemical, biological, radiological, nuclear, or explosive (CBRNE) disasters, fear against invisible hazards causes social panic that often leads to a deterioration in the health of the population.

Sometimes the size of an indirect health impact even surpasses that of a direct impact. For example, after the Fukushima Daiichi nuclear power plant accident in 2011, the limitation of outdoor activities from fear of radiation exposure and other lifestyle changes led to an increase in metabolic syndromes such as hyperlipidaemia[1] and diabetes mellitus[2]. Some researchers estimated that this increase may have even shortened life expectancies to a greater extent than the small amount of radiation exposure caused by the accident[3]. Other health impacts include a decline in physical performance[4], obesity[5, 6], and deterioration of mental status[7] among the evacuees. Considering such situations, preventing indirect impacts is a key to retaining health in disaster areas. Even so, there is a paucity of research about such indirect health impacts and more research is needed to address the chronic conditions after a disaster and how communities can prepare and respond to disasters and public health emergencies.

The SARS-CoV-2 pandemic that started in late 2019 is one of the largest CBRNE disasters of this decade. The virus had killed more than six million by the end of June 2022[8]. In addition, nationwide lockdowns, policies to encourage social distancing, travel restrictions, and voluntary bans of many activities in many countries may have caused severe social disruption and led to lifestyle changes such as alterations in eating habits[9, 10] and a decline in physical activities[11]. Given these situations, experts have raised concerns about an increase in obesity[12, 13] during and after the pandemic. Furthermore, previous research suggested that COVID-19 itself may increase the risk of obesity[14]. Even so, the effect of such social disruption may be heterogeneous. A previous study targeting the population with obesity showed only a limited population were vulnerable to lifestyle changes[15]. Other online surveys have even reported an improvement in body mass index (BMI) and eating habits among some groups of people[16, 17]. However, as these studies targeted the relatively younger population, there is a limitation in the generalizability of the findings. Therefore, a nation-wide survey is needed to understand the size and nature of the risk factors of the indirect impact of the pandemic.

The “Continuing survey on mental and physical health during the COVID-19 pandemic” is a nationwide longitudinal online survey carried out by the Research Institute of Economy, Trade and Industry, Japan (RIETI), Japan. The current study used this data to analyse time trends and risk factors for exercise habit and obesity in Japan So far as well as attitudes toward vaccination[18] and infection avoidance behaviour[19]. The results will provide additional insight on the health impact of a pandemic and therefore will provide clues to developing effective disaster mitigation plans for future CBRNE disasters.

## Materials and Methods

The detailed method of data collection is described in our previous report[18, 19]. In short, nation-wide online questionnaires were conducted five times, that is, October 2020, and January, April, July and October 2021.

### Target population

The participants were Japanese people aged 18–74 years living in Japan who were selected from the database so that their demographic composition ratios of sex, age, and distribution of residential prefectures matched the population estimates of the Statistics Bureau of Japan (final estimates, May 2020). The number of the participants aimed at 2000, according to the eligibility of our fund.

### Data collected

The following data were collected.

- Background information: sex, age group, pre-existing conditions, marital status, yearly income, height, weight, and exercise habit before the COVID-19 pandemic
- Infection status of SARS-CoV-2: past diagnosis, current infection, or no infection
- Activities to avoid the virus: avoid poorly ventilated places, avoid crowded places, wear a mask, wash hands, disinfect belongings, gargle, change clothes frequently, keep a distance from others, refrain from seeing a doctor, and refrain from going out as much as possible
- Exercise habit: days of exercise per week
- Health status: patient health questionnaire 9 (PHQ-9) for depression status[20], GAD-7 for anxiety[21], and subjective health status on a six-point scale
- Change in economic status compared to the previous questionnaire

### Exclusion criteria

As the online survey was written in Japanese, people who could not read Japanese were excluded. After collection, the data were excluded for individuals who provided seemingly inappropriate answers, including non-existent zip codes, extreme outlying values for height and weight, and controversial answers throughout the five questionnaires such as a difference in age of two years or more. The respondents who took a very short time (less than five minutes) or a very long time (ten hours or more) to answer the survey questions were also excluded.

### Definition of changes in the early and late phases

We defined the period of the first and second questionnaire as the ‘early phase’ and that of the fifth questionnaire as the ‘late phase’ of the pandemic. Changes in habits in the early phase were evaluated by comparing the answers in the first and second questionnaires, while changes in the late phase were evaluated by comparing the answers in the first and fifth questionnaires.

### Definition of exercise habit, obesity and overweight

People who answered that they did not exercise (i.e., 0 per week) were categorised as ‘no exercise habit’. Changes in exercise habit were estimated by calculating the difference in exercise days at the time of each questionnaire compared to that stated in the first questionnaire.

Obesity and overweight were defined as a BMI >30 kg/m^2^ and >25 kg/m^2^, respectively. As the proportion of obesity is not so high among Japanese population, proportion overweight is used as outcome for further analysis. Newly developed overweight was defined as those who were not overweight at the time of the first questionnaire but had become overweight in the following periods.

### Statistical analysis

A change in exercise habit during the early phase was calculated by subtracting the exercise days per week in the second questionnaire from the days in the first questionnaire. A change during the late phase was calculated by subtracting the exercise days per week at the time of the fifth questionnaire from the days in the first questionnaire. The difference between exercise days in the first questionnaire and the following questionnaires were analysed using the paired t-test.

The social and psychological impact of the pandemic in Japan has been reported to be different according to sex [22, 23]. Therefore, the statistical analyses were conducted separately by sex. Differences between sex were compared using the chi-square test.

Factors associated with changes in exercise days per week and risk factors for developing overweight were analysed using a multiple linear regression model. For sensitivity analysis, the analysis was conducted using factors in early phase and later phase.

The statistical analyses were carried out using Stata/SE 16.0 (StataCorp LLC, College Station, TX, USA). *P*-values of < 0.05 were considered to be statistically significant.

### Ethical consideration

All individuals who participated in the study provided online consent for their participation. The present study was conducted with the approval of the ethics committee of Hiramatsu Memorial Hospital (No: 20200925).

## Results

Of the 19,340 participants, 2,698 were excluded due to providing inappropriate or controversial answers. The remaining 16,642 (male, 8,022 and female, 8,620) were included for further analysis. The background of the participants grouped by sex are shown in Table 1.

**Table 1.**
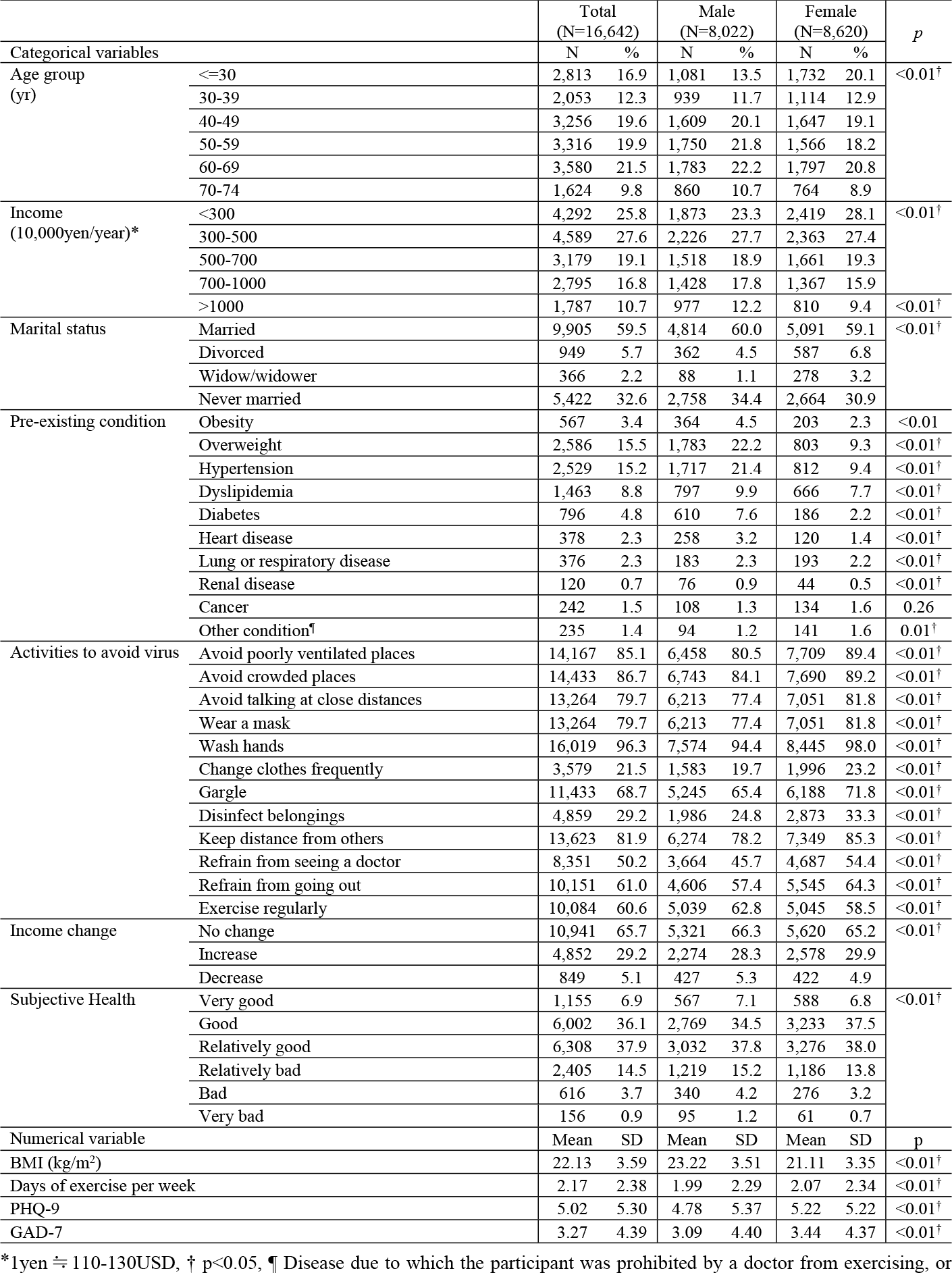
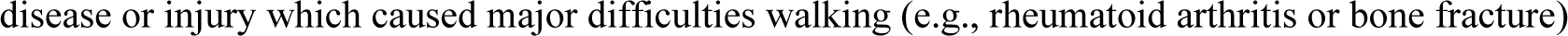
Background of the participants. Differences between males and females were calculated by the chi-squared test for categorical variables and the t-test for numerical variables.

There was a significant difference in all the variables except for the prevalence of cancer. Female participants were more likely to take any infection avoidance behaviour. Male participants had a higher prevalence of exercise habit than females. During the survey, 5,117 (30.7%) of the participants provided at least one missing data or controversial response in the late phase. The background of those with missing data and those with complete data are compared in the S1 table. Younger people, those with lower income levels, and those who never married were more likely to provide essential data.

### Changes in exercise habit

The changes in exercise habits at the time of each questionnaire are shown in Fig 1. The proportion of people who reported less exercise days per week than that at baseline (October 2020) gradually increased with time, while those who reported more exercise days did not change throughout the study period. There was no apparent difference in this trend between males and females. Of the people who reported that they did not exercise at baseline (4,624), 806 (17.4%) reported they had begun to exercise after a year (at the fifth questionnaire). In contrast, 862 (12.6%) of those who reported exercising at baseline (6,841) stopped exercising over the same period. The standard deviations in both males and females increased slightly with time.

**Fig 1.**
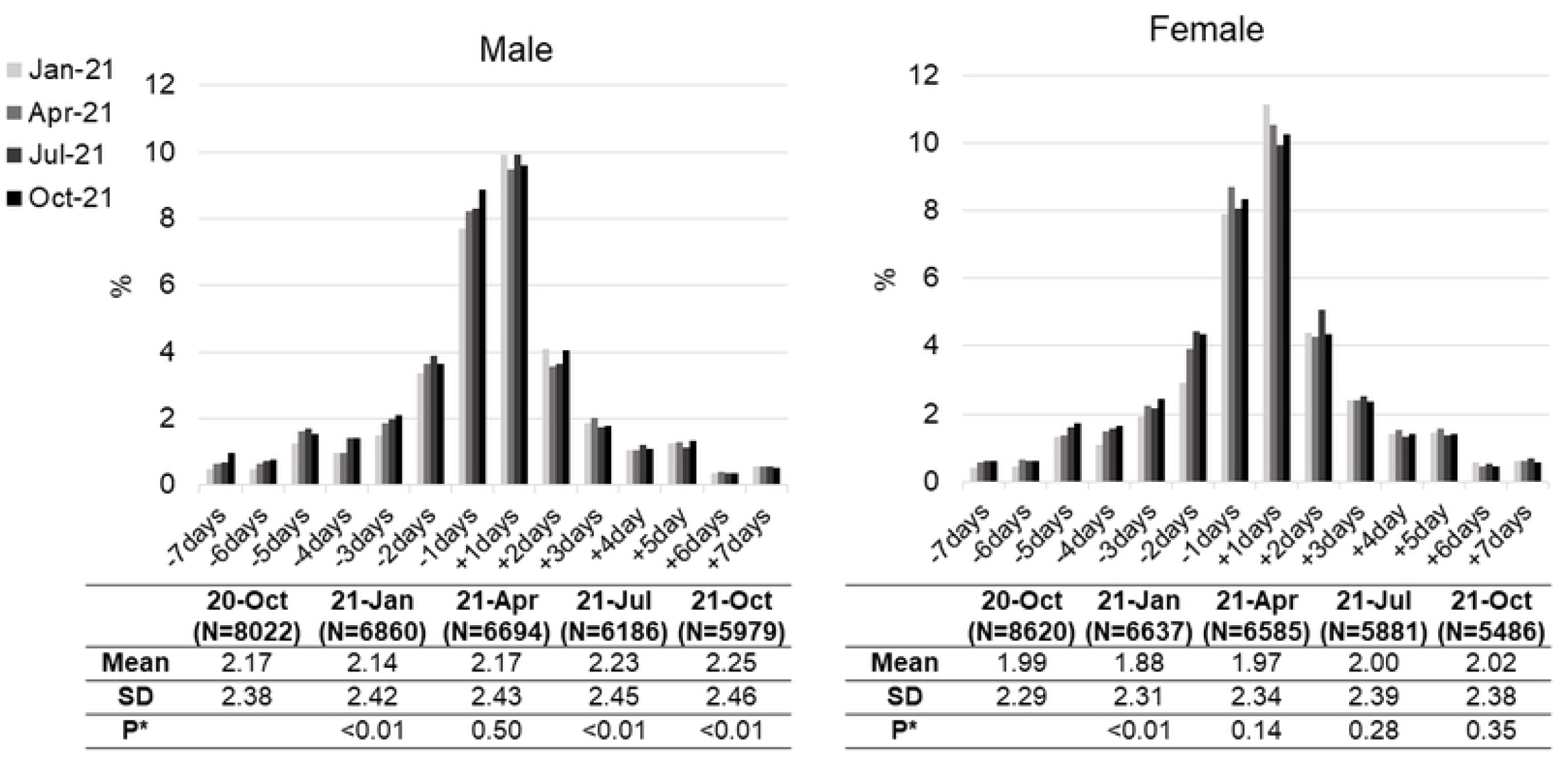
Change in exercise habit (days per week) compared with the first questionnaire (October 2020). Only those who changed the habit are included. * p-values of paired-t test comparing exercise days in each phase with those in October 2020.

To analyse the factors associated with changes in exercise habit, linear regression was conducted using the change in exercise days as the outcome variable. In the early phase of the pandemic (Table 2, left column), a decreased exercise habit was associated with a high income (> 10 million yen per year, equivalent to about 100,000 US dollars per year) in both sex (males, -0.25 days [95% confidence interval -0.44, -0.07]; females, -0.32 days, [-0.51, -0.13]). Elderly females were also associated with decreased exercise days. Having a regular exercise habit at baseline was associated with an increased exercise habit in both sex (males, 1.08 days [0.97, 1.19]; females, 1.28 days [1.17, 1.39]). An increased exercise habit in women was associated positively with PHQ-9 (0.02 (0.01 to 0.04) and negatively with GAD-7 (−0.03 [-0.05, -0.01]), although the size of this effect was small.

**Table 2.**
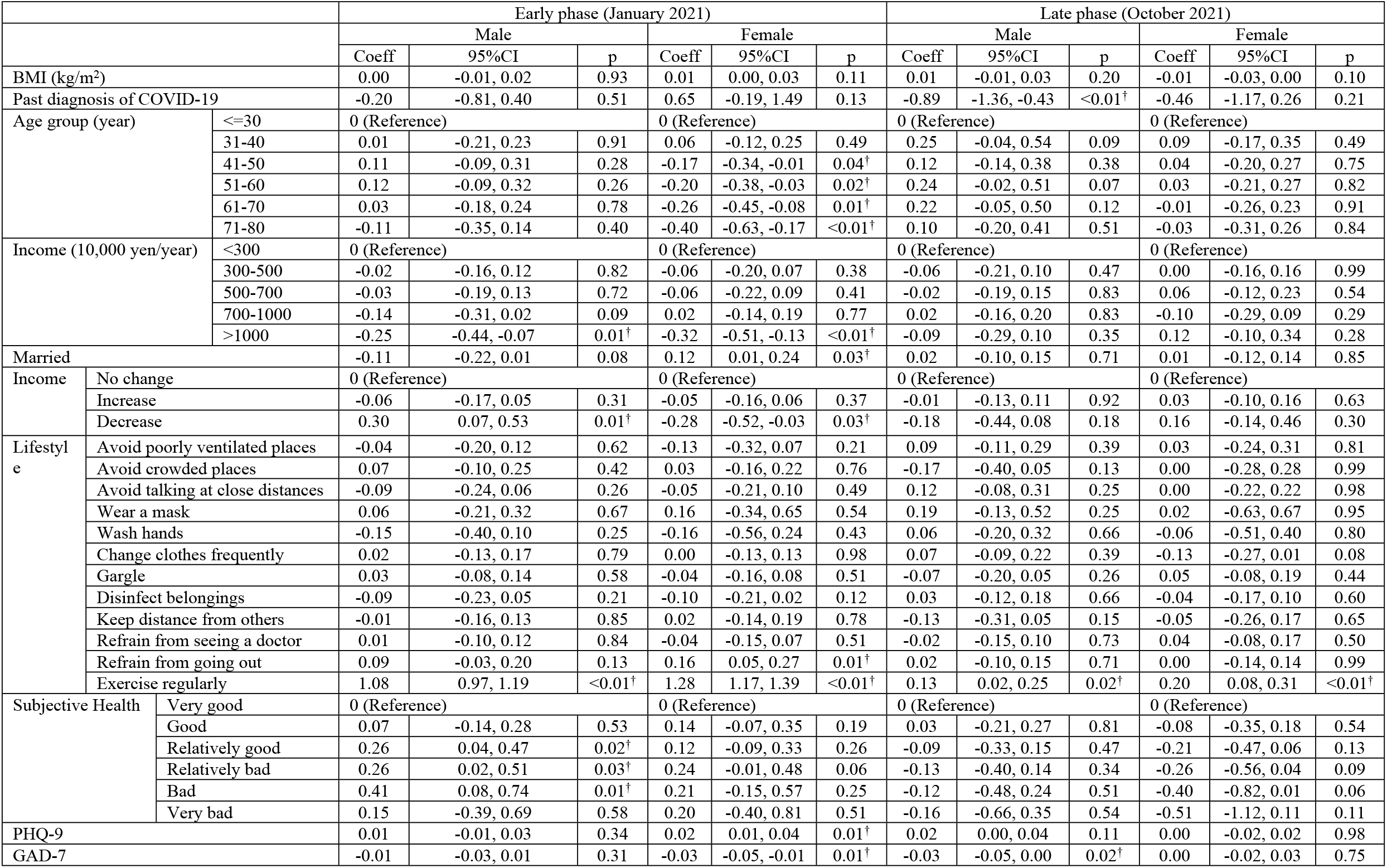
Factors associated with a change in exercise habit in the early phase. Controlled for pre-existing conditions. *10,000yen≒60∼100 USD, † p<0.05

|  |  | Early phase (January 2021) |  |  |  |  |  | Late phase (October 2021) |  |  |  |  |  |
| --- | --- | --- | --- | --- | --- | --- | --- | --- | --- | --- | --- | --- | --- |
|  |  | Male |  |  | Female |  |  | Male |  |  | Female |  |  |
|  |  | Coeff | 95%CI | p | Coeff | 95%CI | p | Coeff | 95%CI | p | Coeff | 95%CI | p |
| BMI (kg/m<sup>2</sup>) |  | 0.00 | -0.01, 0.02 | 0.93 | 0.01 | 0.00, 0.03 | 0.11 | 0.01 | -0.01, 0.03 | 0.20 | -0.01 | -0.03, 0.00 | 0.10 |
| Past diagnosis of COVID-19 |  | -0.20 | -0.81, 0.40 | 0.51 | 0.65 | -0.19, 1.49 | 0.13 | -0.89 | -1.36, -0.43 | <0.01† | -0.46 | -1.17, 0.26 | 0.21 |
| Age group (year) | <=30 | 0 (Reference) |  |  | 0 (Reference) |  |  | 0 (Reference) |  |  | 0 (Reference) |  |  |
|  | 31-40 | 0.01 | -0.21, 0.23 | 0.91 | 0.06 | -0.12, 0.25 | 0.49 | 0.25 | -0.04, 0.54 | 0.09 | 0.09 | -0.17, 0.35 | 0.49 |
|  | 41-50 | 0.11 | -0.09, 0.31 | 0.28 | -0.17 | -0.34, -0.01 | 0.04† | 0.12 | -0.14, 0.38 | 0.38 | 0.04 | -0.20, 0.27 | 0.75 |
|  | 51-60 | 0.12 | -0.09, 0.32 | 0.26 | -0.20 | -0.38, -0.03 | 0.02† | 0.24 | -0.02, 0.51 | 0.07 | 0.03 | -0.21, 0.27 | 0.82 |
|  | 61-70 | 0.03 | -0.18, 0.24 | 0.78 | -0.26 | -0.45, -0.08 | 0.01† | 0.22 | -0.05, 0.50 | 0.12 | -0.01 | -0.26, 0.23 | 0.91 |
|  | 71-80 | -0.11 | -0.35, 0.14 | 0.40 | -0.40 | -0.63, -0.17 | <0.01† | 0.10 | -0.20, 0.41 | 0.51 | -0.03 | -0.31, 0.26 | 0.84 |
| Income (10,000 yen/year) | <300 | 0 (Reference) |  |  | 0 (Reference) |  |  | 0 (Reference) |  |  | 0 (Reference) |  |  |
|  | 300-500 | -0.02 | -0.16, 0.12 | 0.82 | -0.06 | -0.20, 0.07 | 0.38 | -0.06 | -0.21, 0.10 | 0.47 | 0.00 | -0.16, 0.16 | 0.99 |
|  | 500-700 | -0.03 | -0.19, 0.13 | 0.72 | -0.06 | -0.22, 0.09 | 0.41 | -0.02 | -0.19, 0.15 | 0.83 | 0.06 | -0.12, 0.23 | 0.54 |
|  | 700-1000 | -0.14 | -0.31, 0.02 | 0.09 | 0.02 | -0.14, 0.19 | 0.77 | 0.02 | -0.16, 0.20 | 0.83 | -0.10 | -0.29, 0.09 | 0.29 |
|  | >1000 | -0.25 | -0.44, -0.07 | 0.01† | -0.32 | -0.51, -0.13 | <0.01† | -0.09 | -0.29, 0.10 | 0.35 | 0.12 | -0.10, 0.34 | 0.28 |
| Married |  | -0.11 | -0.22, 0.01 | 0.08 | 0.12 | 0.01, 0.24 | 0.03† | 0.02 | -0.10, 0.15 | 0.71 | 0.01 | -0.12, 0.14 | 0.85 |
| Income | No change | 0 (Reference) |  |  | 0 (Reference) |  |  | 0 (Reference) |  |  | 0 (Reference) |  |  |
|  | Increase | -0.06 | -0.17, 0.05 | 0.31 | -0.05 | -0.16, 0.06 | 0.37 | -0.01 | -0.13, 0.11 | 0.92 | 0.03 | -0.10, 0.16 | 0.63 |
|  | Decrease | 0.30 | 0.07, 0.53 | 0.01† | -0.28 | -0.52, -0.03 | 0.03† | -0.18 | -0.44, 0.08 | 0.18 | 0.16 | -0.14, 0.46 | 0.30 |
| Lifestyle | Avoid poorly ventilated places | -0.04 | -0.20, 0.12 | 0.62 | -0.13 | -0.32, 0.07 | 0.21 | 0.09 | -0.11, 0.29 | 0.39 | 0.03 | -0.24, 0.31 | 0.81 |
|  | Avoid crowded places | 0.07 | -0.10, 0.25 | 0.42 | 0.03 | -0.16, 0.22 | 0.76 | -0.17 | -0.40, 0.05 | 0.13 | 0.00 | -0.28, 0.28 | 0.99 |
|  | Avoid talking at close distances | -0.09 | -0.24, 0.06 | 0.26 | -0.05 | -0.21, 0.10 | 0.49 | 0.12 | -0.08, 0.31 | 0.25 | 0.00 | -0.22, 0.22 | 0.98 |
|  | Wear a mask | 0.06 | -0.21, 0.32 | 0.67 | 0.16 | -0.34, 0.65 | 0.54 | 0.19 | -0.13, 0.52 | 0.25 | 0.02 | -0.63, 0.67 | 0.95 |
|  | Wash hands | -0.15 | -0.40, 0.10 | 0.25 | -0.16 | -0.56, 0.24 | 0.43 | 0.06 | -0.20, 0.32 | 0.66 | -0.06 | -0.51, 0.40 | 0.80 |
|  | Change clothes frequently | 0.02 | -0.13, 0.17 | 0.79 | 0.00 | -0.13, 0.13 | 0.98 | 0.07 | -0.09, 0.22 | 0.39 | -0.13 | -0.27, 0.01 | 0.08 |
|  | Gargle | 0.03 | -0.08, 0.14 | 0.58 | -0.04 | -0.16, 0.08 | 0.51 | -0.07 | -0.20, 0.05 | 0.26 | 0.05 | -0.08, 0.19 | 0.44 |
|  | Disinfect belongings | -0.09 | -0.23, 0.05 | 0.21 | -0.10 | -0.21, 0.02 | 0.12 | 0.03 | -0.12, 0.18 | 0.66 | -0.04 | -0.17, 0.10 | 0.60 |
|  | Keep distance from others | -0.01 | -0.16, 0.13 | 0.85 | 0.02 | -0.14, 0.19 | 0.78 | -0.13 | -0.31, 0.05 | 0.15 | -0.05 | -0.26, 0.17 | 0.65 |
|  | Refrain from seeing a doctor | 0.01 | -0.10, 0.12 | 0.84 | -0.04 | -0.15, 0.07 | 0.51 | -0.02 | -0.15, 0.10 | 0.73 | 0.04 | -0.08, 0.17 | 0.50 |
|  | Refrain from going out | 0.09 | -0.03, 0.20 | 0.13 | 0.16 | 0.05, 0.27 | 0.01† | 0.02 | -0.10, 0.15 | 0.71 | 0.00 | -0.14, 0.14 | 0.99 |
|  | Exercise regularly | 1.08 | 0.97, 1.19 | <0.01† | 1.28 | 1.17, 1.39 | <0.01† | 0.13 | 0.02, 0.25 | 0.02† | 0.20 | 0.08, 0.31 | <0.01† |
| Subjective Health | Very good | 0 (Reference) |  |  | 0 (Reference) |  |  | 0 (Reference) |  |  | 0 (Reference) |  |  |
|  | Good | 0.07 | -0.14, 0.28 | 0.53 | 0.14 | -0.07, 0.35 | 0.19 | 0.03 | -0.21, 0.27 | 0.81 | -0.08 | -0.35, 0.18 | 0.54 |
|  | Relatively good | 0.26 | 0.04, 0.47 | 0.02† | 0.12 | -0.09, 0.33 | 0.26 | -0.09 | -0.33, 0.15 | 0.47 | -0.21 | -0.47, 0.06 | 0.13 |
|  | Relatively bad | 0.26 | 0.02, 0.51 | 0.03† | 0.24 | -0.01, 0.48 | 0.06 | -0.13 | -0.40, 0.14 | 0.34 | -0.26 | -0.56, 0.04 | 0.09 |
|  | Bad | 0.41 | 0.08, 0.74 | 0.01† | 0.21 | -0.15, 0.57 | 0.25 | -0.12 | -0.48, 0.24 | 0.51 | -0.40 | -0.82, 0.01 | 0.06 |
|  | Very bad | 0.15 | -0.39, 0.69 | 0.58 | 0.20 | -0.40, 0.81 | 0.51 | -0.16 | -0.66, 0.35 | 0.54 | -0.51 | -1.12, 0.11 | 0.11 |
| PHQ-9 |  | 0.01 | -0.01, 0.03 | 0.34 | 0.02 | 0.01, 0.04 | 0.01† | 0.02 | 0.00, 0.04 | 0.11 | 0.00 | -0.02, 0.02 | 0.98 |
| GAD-7 |  | -0.01 | -0.03, 0.01 | 0.31 | -0.03 | -0.05, -0.01 | 0.01† | -0.03 | -0.05, 0.00 | 0.02† | 0.00 | -0.02, 0.03 | 0.75 |

In the later phase (Table 2, right column), the diagnosis of a SARS-CoV-2 infection was associated with significantly fewer exercise days in males (−0.89 [-1.36, -0.43]). In both sex, a regular exercise habit at baseline remained associated with increased exercise days (males, 0.13 [0.02, 0.25]; females, 0.20 [0.08, 0.31]). In the later phase, mental and physical status had no significant effect on the changes in exercise habit.

### Change in BMI and proportion of obesity/overweight

Another impact caused by the pandemic might be an increase in body weight. The change in BMI between October 2020 and October 2021 are plotted in S1 figure. As some people showed decreased BMI, just calculating mean BMI may not reflect the status of overweight. Therefore, the proportion of obesity/overweight and newly developed overweight as well as mean BMI at each time period are shown in Table 3.

**Table 3.** Fluctuations in body mass index (BMI), proportion of obesity/overweight, and proportion of newly-developed obesity/overweight. For BMI, the values at each time point were compared with those at baseline (October 2020) using the paired t-test.

|  |  | Oct-20 |  | Jan-21 |  |  | Apr-21 |  |  | Jul-21 |  |  | Oct-21 |  |  |
| --- | --- | --- | --- | --- | --- | --- | --- | --- | --- | --- | --- | --- | --- | --- | --- |
|  |  | Mean | SD | Mean | SD | p | Mean | SD | p | Mean | SD | p | Mean | SD | p |
| BMI (kg/m<sup>2</sup>) | Male | 23.22 | 3.54 | 23.41 | 3.99 | <0.01 | 23.39 | 3.79 | <0.01 | 23.34 | 3.78 | 0.07 | 23.34 | 3.74 | 0.08 |
|  | Female | 21.13 | 3.49 | 21.20 | 3.91 | <0.01 | 21.18 | 3.79 | <0.01 | 21.14 | 3.81 | <0.01 | 21.16 | 4.03 | <0.01 |
| Obesity (%) | Male | 4.5 |  | 4.1 |  |  | 4.3 |  |  | 4.2 |  |  | 4.0 |  |  |
|  | Female | 2.4 |  | 2.3 |  |  | 1.9 |  |  | 1.9 |  |  | 2.0 |  |  |
| Newly developed obesity (%) | Male | Base |  | 0.4 |  |  | 0.5 |  |  | 0.6 |  |  | 0.5 |  |  |
|  | Female | Base |  | 0.4 |  |  | 0.3 |  |  | 0.3 |  |  | 0.4 |  |  |
| Overweight (%) | Male | 22.2 |  | 26.6 |  |  | 26.2 |  |  | 25.9 |  |  | 26.1 |  |  |
|  | Female | 9.3 |  | 10.8 |  |  | 10.7 |  |  | 10.6 |  |  | 10.5 |  |  |
| Newly developed overweight (%) | Male | Base |  | 6.9 |  |  | 6.8 |  |  | 6.9 |  |  | 7.2 |  |  |
|  | Female | Base |  | 2.7 |  |  | 2.9 |  |  | 3.0 |  |  | 2.9 |  |  |

Interestingly, the proportions of obesity seem slightly decreased over time, while the proportion of overweight and mean BMI increased in both sex. The standard deviation for BMI also increased over time in females. In addition, the proportion of newly developed obesity among male also increased during the first four questionnaire. The increase in the BMI and the proportion of overweight was marked in the early phase (mean BMI from 23.22 to 23.41 in males and from 21.13 to 21.20 in females; proportion of overweight from 22.2% to 26.6% in males and from 9.3% to 10.8% in females). In the later phase, the change became less marked but remained statistically significant in females.

### Risk factors for developing overweight

As the proportion of obesity was too small to conduct further analysis, factors associated with the development of overweight were determined by multiple logistic regression. In the early phase (January 2021) of the pandemic, the risk of developing overweight was significantly higher in middle-aged males (31-70 years old) and elderly females (71-80 years old) (Table 4, left column). Males who were married were more likely to develop overweight (odds ratio [OR] 1.61 [1.20, 2.16]), although this change was not observed in married females. On the other hand, females who frequently changed their clothes to prevent a COVID-19 infection (OR 1.68 [1.69, 2.60]) or those with a very bad subjective health condition were more likely to develop overweight. An increase in income was also associated with the development of overweight in females (OR 1.54 [1.08, 2.20]), but not in males (OR 0.78 [0.60, 1.01]).

**Table 4.**
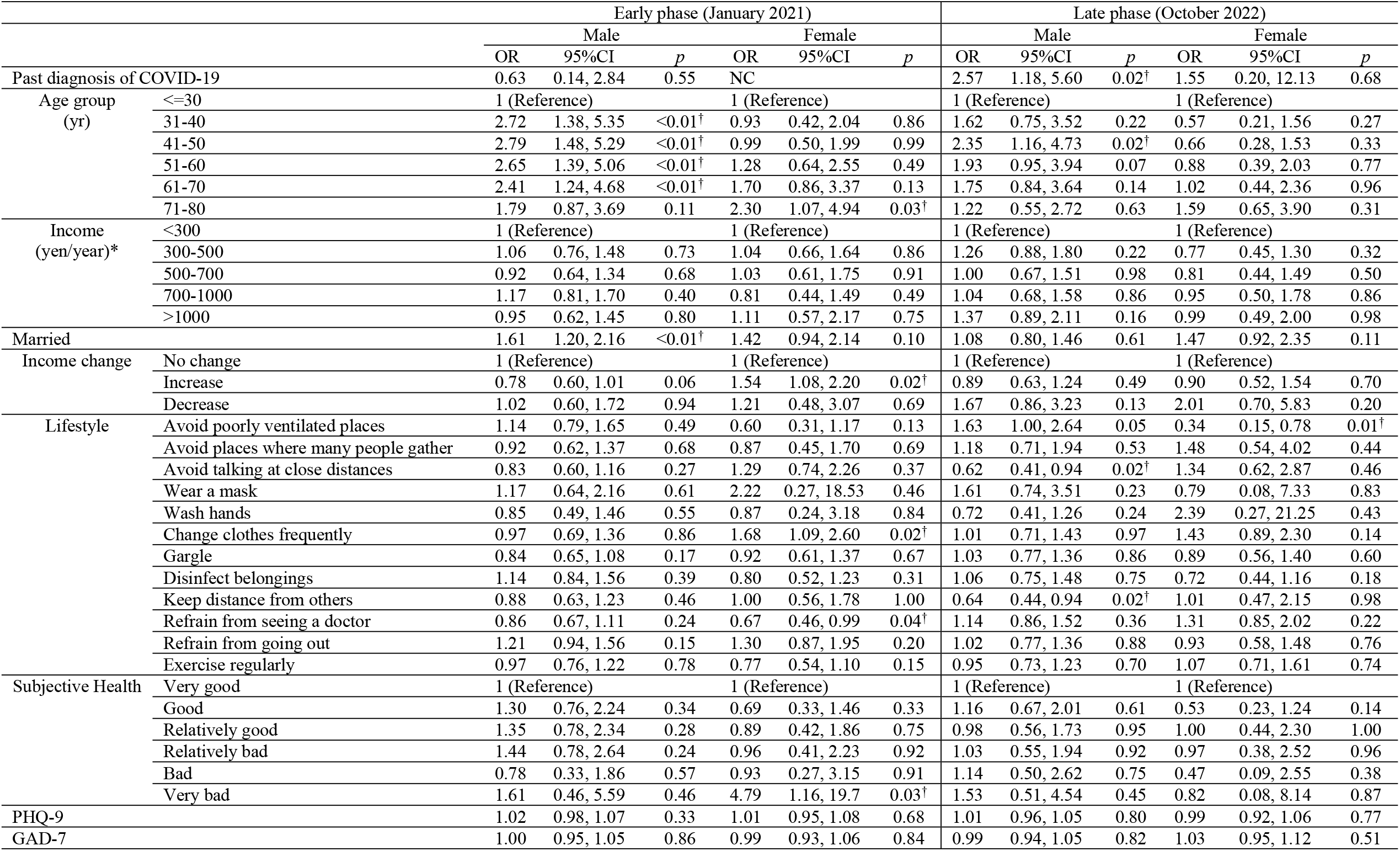
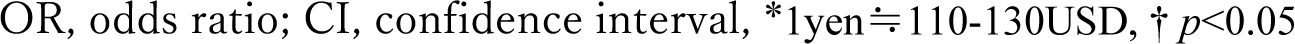
Odds ratios for newly developed overweight in the early and late phases of the pandemic grouped by sex. Controlled for pre-existing conditions.

|  |  | Early phase (January 2021) |  |  |  |  |  | Late phase (October 2022) |  |  |  |  |  |
| --- | --- | --- | --- | --- | --- | --- | --- | --- | --- | --- | --- | --- | --- |
|  |  | Male |  |  | Female |  |  | Male |  |  | Female |  |  |
|  |  | OR | 95%CI | p | OR | 95%CI | p | OR | 95%CI | p | OR | 95%CI | p |
| Past diagnosis of COVID-19 |  | 0.63 | 0.14, 2.84 | 0.55 | NC |  |  | 2.57 | 1.18, 5.60 | 0.02<sup>†</sup> | 1.55 | 0.20, 12.13 | 0.68 |
| Age group (yr) | ≤30 | 1 (Reference) |  |  | 1 (Reference) |  |  | 1 (Reference) |  |  | 1 (Reference) |  |  |
|  | 31-40 | 2.72 | 1.38, 5.35 | <0.01<sup>†</sup> | 0.93 | 0.42, 2.04 | 0.86 | 1.62 | 0.75, 3.52 | 0.22 | 0.57 | 0.21, 1.56 | 0.27 |
|  | 41-50 | 2.79 | 1.48, 5.29 | <0.01<sup>†</sup> | 0.99 | 0.50, 1.99 | 0.99 | 2.35 | 1.16, 4.73 | 0.02<sup>†</sup> | 0.66 | 0.28, 1.53 | 0.33 |
|  | 51-60 | 2.65 | 1.39, 5.06 | <0.01<sup>†</sup> | 1.28 | 0.64, 2.55 | 0.49 | 1.93 | 0.95, 3.94 | 0.07 | 0.88 | 0.39, 2.03 | 0.77 |
|  | 61-70 | 2.41 | 1.24, 4.68 | <0.01<sup>†</sup> | 1.70 | 0.86, 3.37 | 0.13 | 1.75 | 0.84, 3.64 | 0.14 | 1.02 | 0.44, 2.36 | 0.96 |
|  | 71-80 | 1.79 | 0.87, 3.69 | 0.11 | 2.30 | 1.07, 4.94 | 0.03<sup>†</sup> | 1.22 | 0.55, 2.72 | 0.63 | 1.59 | 0.65, 3.90 | 0.31 |
| Income (yen/year)* | <300 | 1 (Reference) |  |  | 1 (Reference) |  |  | 1 (Reference) |  |  | 1 (Reference) |  |  |
|  | 300-500 | 1.06 | 0.76, 1.48 | 0.73 | 1.04 | 0.66, 1.64 | 0.86 | 1.26 | 0.88, 1.80 | 0.22 | 0.77 | 0.45, 1.30 | 0.32 |
|  | 500-700 | 0.92 | 0.64, 1.34 | 0.68 | 1.03 | 0.61, 1.75 | 0.91 | 1.00 | 0.67, 1.51 | 0.98 | 0.81 | 0.44, 1.49 | 0.50 |
|  | 700-1000 | 1.17 | 0.81, 1.70 | 0.40 | 0.81 | 0.44, 1.49 | 0.49 | 1.04 | 0.68, 1.58 | 0.86 | 0.95 | 0.50, 1.78 | 0.86 |
|  | >1000 | 0.95 | 0.62, 1.45 | 0.80 | 1.11 | 0.57, 2.17 | 0.75 | 1.37 | 0.89, 2.11 | 0.16 | 0.99 | 0.49, 2.00 | 0.98 |
| Married |  | 1.61 | 1.20, 2.16 | <0.01<sup>†</sup> | 1.42 | 0.94, 2.14 | 0.10 | 1.08 | 0.80, 1.46 | 0.61 | 1.47 | 0.92, 2.35 | 0.11 |
| Income change | No change | 1 (Reference) |  |  | 1 (Reference) |  |  | 1 (Reference) |  |  | 1 (Reference) |  |  |
|  | Increase | 0.78 | 0.60, 1.01 | 0.06 | 1.54 | 1.08, 2.20 | 0.02<sup>†</sup> | 0.89 | 0.63, 1.24 | 0.49 | 0.90 | 0.52, 1.54 | 0.70 |
|  | Decrease | 1.02 | 0.60, 1.72 | 0.94 | 1.21 | 0.48, 3.07 | 0.69 | 1.67 | 0.86, 3.23 | 0.13 | 2.01 | 0.70, 5.83 | 0.20 |
| Lifestyle | Avoid poorly ventilated places | 1.14 | 0.79, 1.65 | 0.49 | 0.60 | 0.31, 1.17 | 0.13 | 1.63 | 1.00, 2.64 | 0.05 | 0.34 | 0.15, 0.78 | 0.01<sup>†</sup> |
|  | Avoid places where many people gather | 0.92 | 0.62, 1.37 | 0.68 | 0.87 | 0.45, 1.70 | 0.69 | 1.18 | 0.71, 1.94 | 0.53 | 1.48 | 0.54, 4.02 | 0.44 |
|  | Avoid talking at close distances | 0.83 | 0.60, 1.16 | 0.27 | 1.29 | 0.74, 2.26 | 0.37 | 0.62 | 0.41, 0.94 | 0.02<sup>†</sup> | 1.34 | 0.62, 2.87 | 0.46 |
|  | Wear a mask | 1.17 | 0.64, 2.16 | 0.61 | 2.22 | 0.27, 18.53 | 0.46 | 1.61 | 0.74, 3.51 | 0.23 | 0.79 | 0.08, 7.33 | 0.83 |
|  | Wash hands | 0.85 | 0.49, 1.46 | 0.55 | 0.87 | 0.24, 3.18 | 0.84 | 0.72 | 0.41, 1.26 | 0.24 | 2.39 | 0.27, 21.25 | 0.43 |
|  | Change clothes frequently | 0.97 | 0.69, 1.36 | 0.86 | 1.68 | 1.09, 2.60 | 0.02<sup>†</sup> | 1.01 | 0.71, 1.43 | 0.97 | 1.43 | 0.89, 2.30 | 0.14 |
|  | Gargle | 0.84 | 0.65, 1.08 | 0.17 | 0.92 | 0.61, 1.37 | 0.67 | 1.03 | 0.77, 1.36 | 0.86 | 0.89 | 0.56, 1.40 | 0.60 |
|  | Disinfect belongings | 1.14 | 0.84, 1.56 | 0.39 | 0.80 | 0.52, 1.23 | 0.31 | 1.06 | 0.75, 1.48 | 0.75 | 0.72 | 0.44, 1.16 | 0.18 |
|  | Keep distance from others | 0.88 | 0.63, 1.23 | 0.46 | 1.00 | 0.56, 1.78 | 1.00 | 0.64 | 0.44, 0.94 | 0.02<sup>†</sup> | 1.01 | 0.47, 2.15 | 0.98 |
|  | Refrain from seeing a doctor | 0.86 | 0.67, 1.11 | 0.24 | 0.67 | 0.46, 0.99 | 0.04<sup>†</sup> | 1.14 | 0.86, 1.52 | 0.36 | 1.31 | 0.85, 2.02 | 0.22 |
|  | Refrain from going out | 1.21 | 0.94, 1.56 | 0.15 | 1.30 | 0.87, 1.95 | 0.20 | 1.02 | 0.77, 1.36 | 0.88 | 0.93 | 0.58, 1.48 | 0.76 |
|  | Exercise regularly | 0.97 | 0.76, 1.22 | 0.78 | 0.77 | 0.54, 1.10 | 0.15 | 0.95 | 0.73, 1.23 | 0.70 | 1.07 | 0.71, 1.61 | 0.74 |
| Subjective Health | Very good | 1 (Reference) |  |  | 1 (Reference) |  |  | 1 (Reference) |  |  | 1 (Reference) |  |  |
|  | Good | 1.30 | 0.76, 2.24 | 0.34 | 0.69 | 0.33, 1.46 | 0.33 | 1.16 | 0.67, 2.01 | 0.61 | 0.53 | 0.23, 1.24 | 0.14 |
|  | Relatively good | 1.35 | 0.78, 2.34 | 0.28 | 0.89 | 0.42, 1.86 | 0.75 | 0.98 | 0.56, 1.73 | 0.95 | 1.00 | 0.44, 2.30 | 1.00 |
|  | Relatively bad | 1.44 | 0.78, 2.64 | 0.24 | 0.96 | 0.41, 2.23 | 0.92 | 1.03 | 0.55, 1.94 | 0.92 | 0.97 | 0.38, 2.52 | 0.96 |
|  | Bad | 0.78 | 0.33, 1.86 | 0.57 | 0.93 | 0.27, 3.15 | 0.91 | 1.14 | 0.50, 2.62 | 0.75 | 0.47 | 0.09, 2.55 | 0.38 |
|  | Very bad | 1.61 | 0.46, 5.59 | 0.46 | 4.79 | 1.16, 19.7 | 0.03<sup>†</sup> | 1.53 | 0.51, 4.54 | 0.45 | 0.82 | 0.08, 8.14 | 0.87 |
| PHQ-9 |  | 1.02 | 0.98, 1.07 | 0.33 | 1.01 | 0.95, 1.08 | 0.68 | 1.01 | 0.96, 1.05 | 0.80 | 0.99 | 0.92, 1.06 | 0.77 |
| GAD-7 |  | 1.00 | 0.95, 1.05 | 0.86 | 0.99 | 0.93, 1.06 | 0.84 | 0.99 | 0.94, 1.05 | 0.82 | 1.03 | 0.95, 1.12 | 0.51 |

OR, odds ratio; CI, confidence interval, \*1yen $\doteq$ 110-130USD, † $p<0.05$

In the late phase (October 2021) (Table 4, right column), males in the age group of 41-50 yr constantly showed a higher risk of developing overweight (OR 2.35 [1.16, 4.73]). Interestingly, males who were diagnosed with a SARS-CoV-2 infection were also more likely to develop overweight (OR 2.57 [1.18, 5.60]). Avoiding talking at close distances (OR 0.62 [0.41, 0.94]) and keeping distance from others (OR 0.64 [0.44, 0.94]) were also associated significantly with a lower risk of developing overweight in males. In females, avoiding poorly ventilated place was associated with a lower risk of developing overweight (OR 0.34 [0.15, 0.78]).

### Long-term impact of the conditions in the early phase of the pandemic on the onset of overweight

Assuming that overweight in the late phase (October 2021) was affected by factors in the early phase (October 2020), further analysis was carried out on the association between overweight in the late phase and lifestyle factors in the early phase as a sensitivity analysis (S2 table).

In males, infection with the SARS-CoV-2 in the early phase correlated significantly with the development of overweight in the late phase (OR 3.01 [1.27, 7.13]), while those who experienced a decrease in income showed a lower risk (OR 0.73 [0.54, 0.97]). On the other hand, females whose income decreased in the early phase were more likely to show overweight in the late phase (OR 1.75 [1.18, 2.61]). Although not statistically significant, there was a trend that females who had a worse subjective health score in the early phase were more likely to show a higher risk of prolonged overweight. Exercise habit was not associated with the risk of developing overweight in any of the analyses.

## Discussion

This study is a novel nationwide longitudinal research on exercise habits and overweight risks in Japan during the COVID-19 pandemic. The study showed a trend of a decrease in exercise habit and increase in overweight among a group of the population, which suggested the COVID-19 pandemic has had a strong negative impact associated with a restriction of social activities. Even so, our research also showed the proportion of obesity was rather decreased during the period, suggesting the impact was heterogeneous, which was also consistent with previous study [16, 17]. This may mean that not general but targeted intervention might be required to prevent the impact of the disaster on obesity-related health outcomes. In addition, our research revealed that risk factors for immobility and overweight were different, and therefore intervention to prevent these two health problems might be considered independently.

### Older female as a vulnerable population to the COVID-19 pandemic

Importantly, elderly females appeared to be at a higher risk of both immobility and overweight in the early phase of the pandemic. These risks also correlated with worse subjective health in females. These results suggest that the trend might be partly due to fear against COVID-19, which has been reported to be higher in females than in males [24]. In addition, the elderly population might be more vulnerable to biased reports by mass media [25] and the infodemic. Therefore, it is possible that the infodemic and other biased information exacerbated fear against COVID-19 in elderly females. However, this fear may be decreased by fact-checking [24]. Indeed, in other disasters such as the Fukushima nuclear accident, public communication through Fukushima health management surveys was effective for reducing the anxiety among residents [26]. Therefore, in future disasters, appropriate intervention in the acute phase may need to include providing the population with scientific-based information as well as information about self-management and psychological first aid targeting the elderly population.

### Bipolarization of exercise habit

In addition, as our study showed that people who already had an exercise habit were more likely to increase their exercise, improving this pre-condition by installing exercise habits before the pandemic in high-risk groups might be another strategy for disaster preparation.

Interestingly, our study showed that a high income (>10 million yen per year) was associated significantly with decreased exercise habits. This may mean that people engaged in administrative work or work with greater responsibility were overwhelmed by their duty during the pandemic, leading to a decrease in their exercise habit. This may also explain why males who experienced decreased income were more likely to increase their exercise habit. In other words, workload and exercise times were a trade-off in males. On the other hand, females who experienced decreased income were more likely to also decrease their exercise, possibly because those who left their jobs did so due to increased house work[27]. Therefore, exercise times in females were not a trade-off for a reduction in income.

### Concern about the impact of overweight on long-term health conditions

In addition to immobility, obesity is one of the major concerns after a huge disaster, especially among evacuees [6, 28]. The present research study revealed that about 6% of non-obese males and 3% of non-obese females developed overweight during the period of the pandemic. As there was a group of people whose BMI decreased, the net increase in the proportion of overweight was about 4% in males and 1% in females. Above all, middle-aged males were at higher risks of overweight in both the early and late phase of the pandemic. Considering that increased BMI in middle-age causes loss of life expectancy by 5-13 years [29], this indirect impact of the pandemic should not be ignored. To prevent overweight-related disaster impact, intervention in the high-risk population is essential.

#### Risk of overweight among males

For males, the diagnosis of COVID-19 was associated significantly with the development of overweight. As the diagnosis also correlated with a decrease in exercise habits, this increase in overweight may have been due to lack of exercise. However, there was no significant difference in exercise days between those who were diagnosed with a SARS-CoV-2 infection and those who were not (data not shown). Another possible reason is that post-COVID syndromes such as post-traumatic disorders, depression, and chronic fatigue may lead to inactivity and increased the risk of overweight [14]. Some experts consider rehabilitation in the recovery phase of COVID-19 should include not only respiratory and cardiovascular rehabilitation but also muscle training and psychological supports[30]. Such interventions may need to be applied for those whose symptoms were less severe. However, to date there are no guidelines regarding interventions for patients who were not hospitalised. An effort to reduce the indirect and prolonged health impacts caused by the SARS-CoV-2 pandemic may need to target this population.

#### Risk of overweight among females

Our research also revealed elderly females were at higher risk of developing overweight in the early phase compared with other age groups. A previous study reported homemakers were more likely to gain body weight [15], which was consistent with our findings. The factors causing overweight in elderly people include a decrease in time spent for outings due to geographically isolated conditions of temporary housings [31] and prolonged post-traumatic stress disorders (PTSD) [32]. During the COVID-19 pandemic, people stayed at home for a longer time, which might have caused a similar condition to long-term evacuation such as less outings and higher mental stress. Overweight in elderly women may have a marked health impact on society because overweight in this population group is a significant risk factor for immobility and frailty, which may lead to bone fracture or a bed-ridden state [33, 34]. Therefore, immediate intervention might have been needed to target this group of people in the early phase of the pandemic. For females, the development of overweight was associated with seemingly excessive reactions against SARS-CoV-2, such as changing clothes frequently. As bad subjective health status is associated with the risk of developing overweight, anxiety might have also been a risk factor.

To prevent lifestyle diseases, interventions by health professionals are not sufficient. In addition, the health system is often severely compromised in the affected areas due to overwhelming demand, evacuation of healthcare workers[35], diversion of resources, and closure of health facilities [36]. Therefore, self-management such as regular exercise and weight control is a key to disaster mitigation.

### Limitations

This study had several limitations. First, the study relied solely on participant responses and therefore we could not avoid false answers even after excluding those that were apparently controversial. Second, although the participants were matched to the national demographic background there remains selection bias of the participants. For example, individuals who could not read Japanese and those who could not use the internet were excluded. In addition, individuals with a history of infection could have more actively sought to participate in our study because of their increased interest in the significance and content of this online survey, causing an upward bias in participation of this type of subject. Third, about one-third of the participants dropped out during the survey period. As there were some significant differences between those with missing data and those with complete data (S1 table), these numbers may have affected the generalizability of the results. Finally, causal relationships cannot be proved by this survey. For example, it is not clear whether newly developed overweight increased the risk of COVID-19 infection or vice-versa. However, by using the factors in the early phase as explanatory variables and newly developed overweight as an outcome variable, this limitation could be partially overcome. Therefore, despite these limitations, our research provided sufficient generalizability compared to other studies.

## Conclusion

This study analysed the impact of the COVID-19 pandemic on exercise habit and development of overweight in the Japanese population. Risk factors for these conditions are shown to be different between sex. Our results suggest that early intervention for elderly women such as provision of information and mental care, and long-term intervention including physical and mental rehabilitation for those who were infected might have been needed. As most CBRNE disasters can cause similar social transformation, intervention to prevent immobility and obesity among high-risk population should be addressed in future disaster preparation/mitigation plans so that we can prevent distant health impact by a disaster.

## Data Availability

As the data was collected by an outsourcing company, the data authorship, data can not be shared publicly.

## Acknowledgement

We would like to thank Dr Kenzo Denda, Hiramatsu Memorial Hospital for his support in ethical considerations of the study.

